# Serum Neurofilament Light Chain Increases in Healthy Postpartum: Is It Subclinical Brain Damage or Neuroplasticity?

**DOI:** 10.1101/2025.11.01.25338915

**Authors:** Antonio Bertolotto, Paola Valentino, Cecilia Irene Bava, Serena Martire, Simona Malucchi, Carola Beltratti, Alessia Di Sapio, Luca Marozio

## Abstract

**Background:** Elevated serum neurofilament light chain (sNfL) levels occur postpartum in women with pre-eclampsia or multiple sclerosis (MS), though definitive central nervous system (CNS) injury remains unestablished. Data on sNfL levels in healthy women with physiological pregnancies (HWwPP) are lacking, whereas numerous MRI and histological studies have demonstrated pregnancy-induced neuroplasticity.

**Methods:** We collected blood samples from 61 HHwPP at four peripartum time points (−1, +1, +2-5, +6-10 days), including nulliparous, primiparous, and multiparous women. Two additional control groups (19 healthy women at 20-180 days postpartum; 43 healthy women remote from delivery) were studied. Serum levels of NfL, Glial Fibrillary Acidic Protein (GFAP), Tubulin Associated Unit (TAU) protein, and Ubiquitin C-terminal Hydrolase L1 (UCHL1) were analyzed.

**Results:** sNfL levels doubled on postpartum day 1 (median 19.84; range 5.02-66.19) versus pre-delivery (8.86; 1.88-34.62) (p<.001), increasing further at 6-10 days (28.18; 7.61-48.83). sNfL decreased significantly with increasing parity (linear mixed effect model: b=−0.74, 95% CI [−0.620, −0.872], p=0.000642). This parity effect was also observed for sUCHL1, sGFAP, and sTAU.

sUCHL1 increased sharply from median of 24.51 (1.58-170.63) before delivery to 153.69 (11.25-971.81) (p<.001) on postpartum day 1, returning to baseline by days 6-10.

**Conclusion:** HWwPP showed significant postpartum increase in sNfL and sUCHL1, most pronounced in the first pregnancy and diminishing with increasing parity. In women with MS, these elevations should not be attributed solely to new CNS lesions. Further studies must clarify whether this increase indicates subclinical CNS damage or reflects pregnancy-induced neuroplasticity.

**What is Already Known:** Increased sNfL levels are considered a marker of nervous system lesion. Elevated postpartum sNfL levels have been reported in most, but not all, women with MS or pre-eclampsia, without clear evidence of CNS injury.

No data are currently available for healthy women with physiological pregnancies (HWwPP).

Pregnancy induces significant neuroplasticity in the CNS

**What This Study Adds:** This study demonstrates a consistent postpartum increase in sNfL and ubiquitin carboxyl-terminal hydrolase L1 (UCHL1) levels in most, but not all, HWwPP. The transient increase of sNfL and sUHCL1 decreases with increasing parity.

**How This Study Might Affect Research, Practice, or Policy:** The postpartum increase in sNfL in women with MS not necessarily indicate new CNS lesions and MS disease activity.

These findings suggest that the elevated postpartum sNfL and UCHL1 levels may reflect a physiological neuroplasticity induced by pregnancy.

This study enables investigation of associations between biomarker levels and postpartum psychiatric disorders.

## Introduction

Elevated serum neurofilament light chain (sNfL) is a biomarker for neuroaxonal injury in multiple sclerosis (MS), amyotrophic lateral sclerosis, neurodegenerative disorders, and acute brain and spinal cord injury.^1^ In MS, sNfL correlates with disease activity, disability, patient outcomes, and therapeutic response.^2-5^

Studies examining sNfL during pregnancy and postpartum in women at risk of pre-eclampsia^6^ and in women with MS (wwMS) treated with alemtuzumab^7^ or natalizumab^8^ consistently found sharp, transient sNfL increases in most patients within days after delivery. Mean sNfL levels were also elevated in wwMS compared to healthy women during the first trimester postpartum, though exact sampling times were not reported.^9^ While this postpartum sNfL increase could reflect CNS injury from new MS lesions or preeclampsia risk,^10-12^ none of the wwMS with elevated sNfL within 8 weeks postpartum showed clinical or radiological disease activity.^7,8^ Similarly, our research group at the Regional Reference Center for Multiple Sclerosis (CRESM), Orbassano, Italy, observed high postpartum sNfL levels without concurrent clinical or MRI activity in 3 of 3 wwMS (unpublished data).

To determine the dynamics of postpartum sNfL elevation and its association with parturition itself, we conducted a longitudinal study in healthy women with physiological pregnancies (HWwPP). To assess potential CNS involvement, we also measured astrocyte damage marker Glial Fibrillary Acidic Protein (GFAP) and neuronal markers Tubulin Associated Unit (TAU) protein and Ubiquitin C-terminal Hydrolase L1 (UCHL1).

## Materials and methods

### Mothers, pregnancies and newborns

A longitudinal study included 61 healthy women with physiological pregnancies (HWwPP) aged 18–42 years with singleton pregnancies who delivered at Sant’Anna University Hospital, Turin, Italy (January 2023–February 2025). Exclusion criteria were: preexisting or gestational diabetes mellitus requiring insulin, chronic hypertension, renal disease, neurological or autoimmune disorders, pregnancy-induced hypertension, uteroplacental dysfunction, and previous preeclampsia, eclampsia, or HELLP syndrome. Deliveries occurred via spontaneous vaginal delivery (n=28, 45.9%) or caesarean section (C-section; n=33, 54.1%). Among C-sections, 31 were elective; two were urgent due to failed labor induction and fetal distress. Twelve participants (19.7%) had minor preexisting conditions (hypothyroidism n=3; thrombocytopenia, prothrombin deficiency, thalassemia minor carrier, thyroid cancer history, hypercholesterolemia, papillitis, psoriasis, congenital heart disease, and growth hormone deficiency, n=1 each). Table 1 presents maternal and pregnancy characteristics: age, ethnicity, pre-pregnancy BMI, parity (0=nulliparous, 1=second pregnancy, ≥2=third/fourth pregnancy), conception mode (spontaneous/medically assisted), delivery mode, gestational age at delivery (all term: 37–41 weeks), postpartum complications, and breastfeeding status (56/61 initiated immediately postpartum; 5 received cabergoline for lactation suppression).

**Table 1.** Demographic and clinical characteristics of mothers, pregnancies and newborns. Abbreviations AGA = appropriate for gestational age; BMI = Boby mass index; C-section = caesarean section; LGA = large for gestational age; SGA = small for gestational age

| **Mothers** |  |  |
| --- | --- | --- |
| Age, years m ± SD, range | | 35.4 ± 4.4 (18-42) |
| Ethnicity n % |  |  |
|  | White | 55 (90.1%) |
|  | North-African | 3 (4.9%) |
|  | Latino | 2 (3.3%) |
|  | Asian | 1 (1.6%) |
| BMI m ± SD, range | | 22.2 ± 3.2 (18-30) |
| **Pregnancies** |  |  |
| Conception, n % |  |  |
|  | Spontaneous | 56 (91.8%) |
|  | Medically Assisted | 5 (8.25%) |
| Parity, n % |  |  |
|  | 0 | 22 (36.1%) |
|  | 1 | 31 (50.8%) |
| | $\geq 2$ | 8 (13.1%) |
| Delivery, n % |  |  |
|  | Vaginal | 28 (45.9%) |
|  | C-section | 33 (54.1%) |
| Epidural anesthesia in vaginal delivery, n (%) |  |  |
|  | Yes | 8 (28.6%) |
|  | No | 20 (71.4%) |
| Partum complications, n % |  |  |
|  | Post-partum hemorrhage | 7 (11.4%) |
|  | Hypertension | 1 (1.6%) |
|  | Fever | 1 (1.6%) |
| Breastfeeding n % |  |  |
|  | Yes | 56 (91.8%) |
|  | No | 5 (8.2%) |
| **Newborns** |  |  |
| Sex, n (%) |  |  |
|  | Male | 31 (50.8%) |
|  | Female | 30 (49.1%) |
| Neonatal weight, g ± SD | | 3344.3 ± 402.9 |
| Gestational week at birth, n (%) |  |  |
|  | 37 weeks | 8 (13.1%) |
|  | 38 weeks | 27 (44.3%) |
|  | 39 weeks | 14 (22.9%) |
|  | 40 weeks | 12 (19.6%) |
| Fetal Biometry |  |  |
|  | SGA | 0 (0%) |
|  | AGA | 55 (90.1%) |
|  | LGA | 6 (9.9%) |
| 5 min Apgar score <7, n % |  | 1 (1.6%) |

Newborn data in Table 1 include: sex, birth weight, gestational age, fetal biometry (third-trimester ultrasound classified as appropriate [AGA, 10th–95th percentile], large [LGA, >95th percentile], or small [SGA, <10th percentile] for gestational age), and 5-minute Apgar score.

### Blood samples

Blood samples were collected from all 61 HWwPP at three timepoints: one day before delivery (−1D; median 15 h, range 1–42 h), one day after delivery (+1D; median 16 h, range 1–33 h), and 2–5 days after delivery (+2-5D; median 63 h, range 34–91 h). A fourth sample was collected from 46 of these women at 6–10 days postpartum (+6-10D; median 206 h, range 120–522 h).

To assess long-term pregnancy effects on sNfL, we collected 19 blood samples from healthy women at 1–6 months postpartum, forming the “late post-pregnancy” (LPP) group.

Baseline sNfL levels were established from 42 healthy, non-pregnant women (N-PW) aged 20–43 years who were either nulligravid or >9 months postpartum. Samples for both LPP and N-PW groups were obtained from the CRESM Biobank, which provides biological samples and data from MS patients and controls.^13^

Blood samples were centrifuged at 3000 × g for 10 min, and serum was stored at −80°C in coded aliquots to prevent freeze-thaw cycles.

### Biomarker quantification

sNFL, GFAP, TAU protein and UCHL1 were quantified using SIMOA Neurology 4-plex assay (Quanterix) on SR-X™ Biomarker Detection System (Quanterix) in the Neurobiology Laboratory of CRESM. Samples were analyzed following manufacturer’s instruction. Assay staff was blinded to women’s clinical characteristics.

### Statistical analysis

Statistical analysis was performed using R version 4.4.1. Normality of distribution and homogeneity of variances were assessed using the Shapiro-Wilk test and Levene’s test, respectively. Descriptive statistics for clinical, demographic, and biological data were calculated using Student’s t-test, Fisher’s exact test, and Mann-Whitney U tests, as appropriate. Differences between groups were evaluated using the Friedman test with Dunn’s post hoc test for longitudinal data, and the Kruskal-Wallis test with Dunn’s post hoc test for comparisons across different categories. Associations between biomarker levels and blood draw timing, parity, and type of birth, including their interactions, were determined using a linear mixed-effects model to account for intra-individual variability. A p-value of <0.05 was considered statistically significant.

## Results

### sNFL levels over time

The mean sNFL in 42 samples of N-PW was 4.79 pg/ml (SD ±1.70, median 4.55 pg/ml, range 1.70 - 9.40); the mean +3SD was 9.88 pg/ml (Figure 1A). The latter value is within the range of healthy controls younger than 40 years as found in our previous study regarding sNFL cut-off determination^14^.

**Figure 1.**
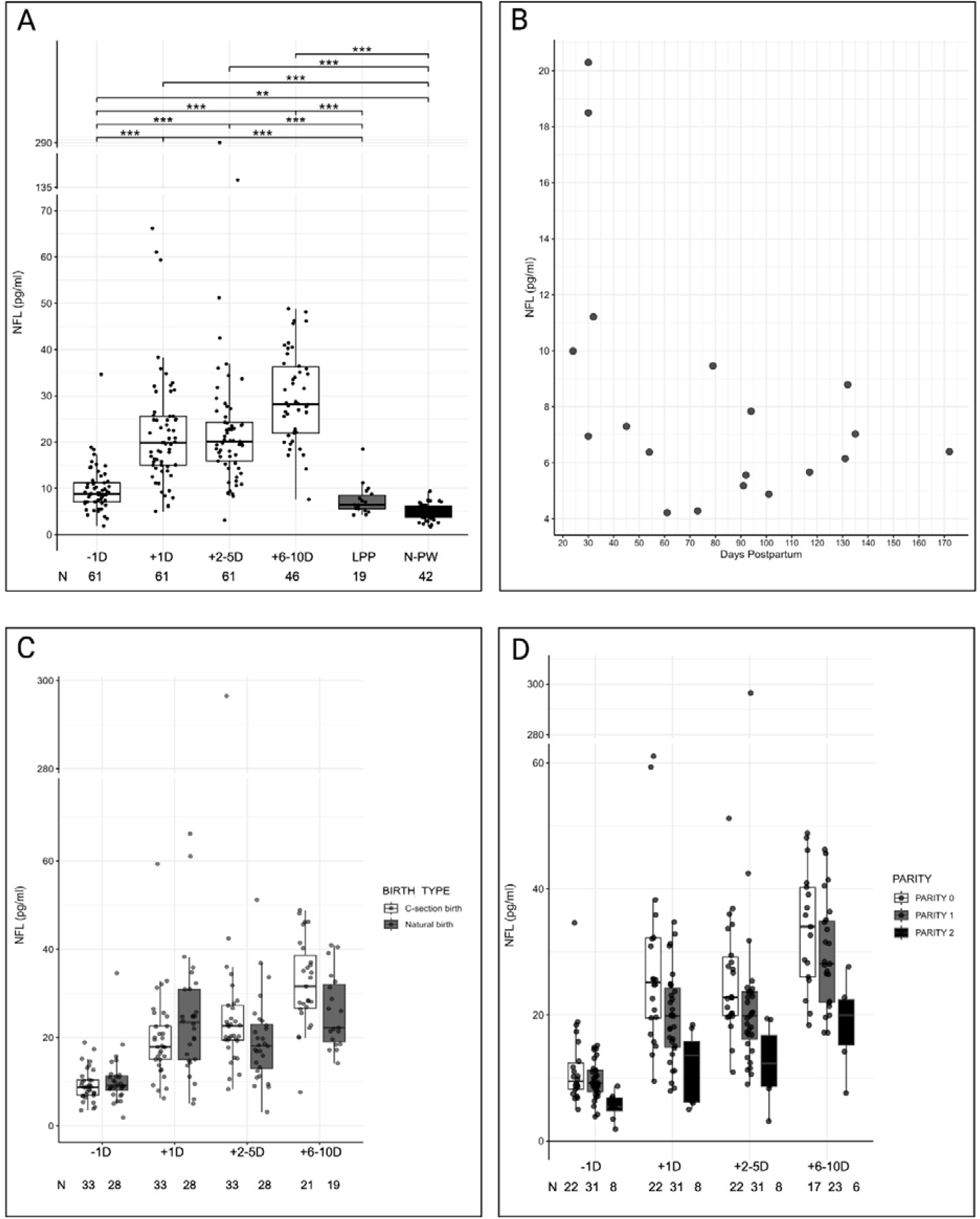
sNfL and pregnancy. -1D: the day before partum; +1D: the day post-partum; +2-5D: 2-5 days post-partum; +6-10D: 6-10 days post-partum; LPP: late post-pregnancy, 20 −180 days post-partum; N-PW: non-pregnant women. Boxes represent interquartile ranges (25th to 75th percentiles), and horizontal lines indicate median values. ** p < 0.001; *** p < 0.0001, (see test for statistical analysis). **A Levels of sNfL pre- and post-partum, 20-180 days post-partum and in non-pregnant women**. **B Level of sNfL in 19 women between 20 and 180 days after delivery** **C sNfL levels pre- and post-partum in vaginal and C-section delivery** **D Impact of parity on sNfL levels**

The levels of sNFL in 61 blood samples collected before delivery had a median value of 8.86 pg/ml (range 1.88-34.60); at the +1D the median sNFL value showed a sharp increased at 19,84 g/ml (range 5.02-66.19), (Figure 1A; Table2).

**Table 2.** Serum biomarkers levels in pre- and post-partum, according to the type of delivery and the parity. Abbreviations −1D: first time-point, the day before partum; +1D: second time-point, the day postpartum; +2-5D: third time-point, 2-5 days post-partum; +6-10D: forth time-point, 6-10 days post-partum; Parity 0 = first pregnancy; Parity 1 = second pregnancy; Parity ≥ 2 = third or fourth pregnancy; sUCHL1 = Ubiquitin C-terminal Hydrolase L1; sGFAP = serum Glial Fibrillary Acidic Protein; sNfL = serum Neurofilaments Light; sTAU = serum Tubulin

|  |  | **sNFL, pg/ml** |  |  |  |
| --- | --- | --- | --- | --- | --- |
| Time-points |  | -1D | +1D | +2-5D | +6-10D |
| All pregnancies | Number | 61 | 61 | 61 | 46 |
|  | Median (range) | 8.86<br>(1.88 - 34.62) | 19.84<br>(5.02 - 66.19) | 20.15<br>(3.14 - 296.48) | 28.18<br>(7.61 - 48.83) |
| Vaginal delivery | Number | 28 | 28 | 28 | 19 |
|  | Median (range) | 9.10<br>(1.88 - 34.62) | 23.41<br>(5.02 - 66.19) | 18.13<br>(3.14 - 51.18) | 22.23<br>(14.19 - 40.91) |
| C-section | Number | 33 | 33 | 33 | 27 |
|  | Median (range) | 8.73<br>(3.49 - 18.88) | 18.13<br>(3.14 - 51.18) | 22.61<br>(8.32 - 296.48) | 31.56<br>(7.61 - 48.83) |
| Parity 0 | Number | 22 | 22 | 22 | 17 |
|  | Median (range) | 9.48<br>(5.02-34.60) | 25.17<br>(9.48-66.19) | 22.78<br>(10.91-51.18) | 34.00<br>(18.38-48.83) |
| Parity 1 | Number | 31 | 31 | 31 | 23 |
|  | Median (range) | 9.19<br>(3.87-15.16) | 19.84<br>(7.93-34.75) | 19.88<br>(8.99-296.48) | 28.11<br>(17.16-46.23) |
| Parity ≥ 2 | Number | 8 | 8 | 8 | 6 |
|  | Median (range) | 5.50<br>(1.88-8.71) | 13.56<br>(5.02-18.42) | 12.30<br>(3.14-19.40) | 19.95<br>(7.61-27.65) |
| **sUCHL1, pg/ml** |  |  |  |  |  |
| All pregnancies | Number | 61 | 61 | 61 | 46 |
|  | Median (range) | 24.51<br>(1.58 - 170.63) | 153.69<br>(11.25 - 971.81) | 39.10<br>(10.16 - 239.35) | 27.74<br>(9.91 - 380.76) |
| **sGFAP, pg/ml** |  |  |  |  |  |
| All pregnancies | Number | 61 | 61 | 61 | 46 |
|  | Median (range) | 85.56<br>(41.77 - 305.10) | 83.51<br>(50.33 - 468.17) | 83.88<br>(40.58 - 298.90) | 81.45<br>(39.85 - 230.14) |
| **sTAU, pg/ml** |  |  |  |  |  |
| All pregnancies | Number | 61 | 61 | 61 | 46 |
|  | Median (range) | 0.69<br>(0.08 - 4.51) | 0.85<br>(0.29 - 15.34) | 0.71<br>(0.25 - 3.18) | 0.75<br>(0.28 - 3.91) |

The levels of sNfL remained elevated at +2-5D and increased further at +6-10D, with median values of 20.15 pg/ml (range: 3.14 – 296.48) and 28.19 pg/ml (range: 7.61 – 48.83), respectively (Figure 1A; Table 2).

The median sNfL level in 19 LPP serum samples collected 20-180 days postpartum was 6.95 pg/ml (range: 4.22–20.29) (Figure 1A). Specifically, 4 of the 5 samples collected between 20 and 40 days postpartum showed a value above 9.88 pg/ml (the mean + 3SD cut-off derived from the N-PW group), while all 14 samples collected between 40 and 180 days postpartum were below this threshold (Figure 1B). Compared to −1D, sNFL levels were significantly higher at times +1D, +2-5D and +6-10D (Friedman’s test, all p<0.0001). Compared to +6-10D, +1D and +2-5D sNFL levels were significantly lower (Friedman’s test, p=0.0022 and p=0.0083, respectively).

All 3 post-delivery time points also showed significantly higher sNfL values compared to both the LPP and N-PW groups (Kruskal-Wallis test, both p<0.0001). At −1D, sNfL levels were not significantly different from the LPP group but were statistically higher than the N-PW group (Kruskal-Wallis test, p=0.0031) (Figure 1A).

The postpartum increase in sNfL was observed in most women, with some notable exceptions. Using 9.88 pg/ml as the upper limit for the N-PW group, sNfL levels above this value were found in 23/61 women (37.7%) at −1D, 54/61 women (88.5%) at +1D, 56/61 women (91.8%) at +2-5D, 45/46 women (97.8%) at +6-10D. One woman (1.6%) had sNfL values below the 9.88 pg/ml threshold in all 4 longitudinal samples. Two other women showed values above the threshold only at the +6-10D time point. Interestingly, all 3 of these women were on their third or fourth pregnancy, with 1 having a C-section and the other 2 a vaginal delivery.

### Levels of NfL in Vaginal and C-section deliveries

Twenty-eight women underwent vaginal delivery and 33 underwent C-section, with no significant age difference between the two groups.

A linear mixed effect model with log-transformed sNFL as dependent variable and accounting for time from delivery, parity, type of birth, with relative interaction with time, and intra-individual variability evidenced that the type of delivery had no statistically significant main effect on sNFL levels (Table 2; Figure 1C).

### Impact of parity on sNfL

Parity was categorized by the number of previous births (term or preterm). The sample included 22 nulliparous women (Parity 0), 31 primiparous (Parity 1), and 8 multiparous (Parity ≥ 2), of whom four were at their third pregnancy (Parity 2) and four at their fourth (Parity 3). Since the latter two groups showed no significant difference in mean sNFL levels (t(28)=1.64,p=0.112), they were combined for analysis.

Thirteen women had miscarriage or abortion: four were nulliparous, five primiparous, and four multiparous. The three parity groups showed no statistically significant differences in age (Kruskal test, p=0.560) or in the distribution of delivery types (Fisher’s exact test, p=0.7714) (Table 3).

**Table 3.** Distribution of participants according to type of delivery and parity. Abbreviations Parity 0 = first pregnancy; Parity 1 = second pregnancy; Parity = 2 = third or fourth pregnancy

| **PARITY** | **AGE –**<br>**median**<br>**years**<br>**(range)** | **Age**<br>**difference**<br>**p-value**<br>**(Kruskal**<br>**Wallis)** | **TOTAL**<br>**deliveries**<br>**(N)** | **NATURA**<br>**L**<br>**delivery**<br>**(N)** | **C-SECTION**<br>**delivery**<br>**(N)** | **Type of**<br>**delivery**<br>**distribution**<br>**p-value**<br>**(Fisher’s**<br>**exact test)** |
| --- | --- | --- | --- | --- | --- | --- |
| 0 | 35<br>(25-42) | 0.560 | 22 | 11 | 11 | 0.7714 |
| 1 | 36<br>(23 - 41) |  | 31 | 13 | 18 |  |
| ≥ 2 | 32<br>(25 - 41) |  | 8 | 4 | 4 |  |

A linear mixed effect model with log-transformed sNFL as dependent variable and accounting for time from delivery, type of birth, parity, with relative interaction with time, and intra-individual variability showed that parity significantly and consistently impact the levels of sNFL in a time-independent fashion, with decreasing levels of sNFL in patients with a history of higher number of previous pregnancies (b = −0.74, 95% CI [(−0.620, −0.872], p=0.000642), (Table 2; Figure 1D).

### Impact of breastfeeding on the levels of sNfL

Since most Italian women initiate breastfeeding immediately postpartum, only five (8.2%) women in our study did not, precluding analysis of breastfeeding’s impact on sNfL levels.

### sUCHL1 Levels over time

Median sUCHL1 in 61 pre-delivery samples (−1D) was 24.51 pg/ml (range: 1.01– 170.63 pg/ml). A sharp, transient peak occurred one day post-delivery (+1D), with levels increasing >6-fold to 153.69 pg/ml (range: 11.25–971.81 pg/ml; Friedman test, −1D vs +1D, p<0.0001) (Table 2, Figure 2B). Levels declined rapidly, reaching 39.15 pg/ml at +2-5D (range: 7.39–239.35 pg/ml; Friedman tests: −1D vs +2-5D, p=0.0033; +1D vs +2-5D, p<0.0001). By +6-10D (n=46), levels returned to 27.74 pg/ml (range: 9.91–380.76 pg/ml), comparable to pre-delivery values (Friedman test, −1D vs +6-10D, ns) but significantly lower than the +1D peak (p<0.0001). Median sUCHL1 in 19 LPP was 22.48 pg/ml (range: 13.78–35.56 pg/ml). While −1D and +6-10D levels did not differ from LPP, both +1D (Kruskal-Wallis test, p<0.0001) and +2-5D (p=0.0019) showed significantly elevated levels (Table 2, Figure 2B).

**Figure 2.**
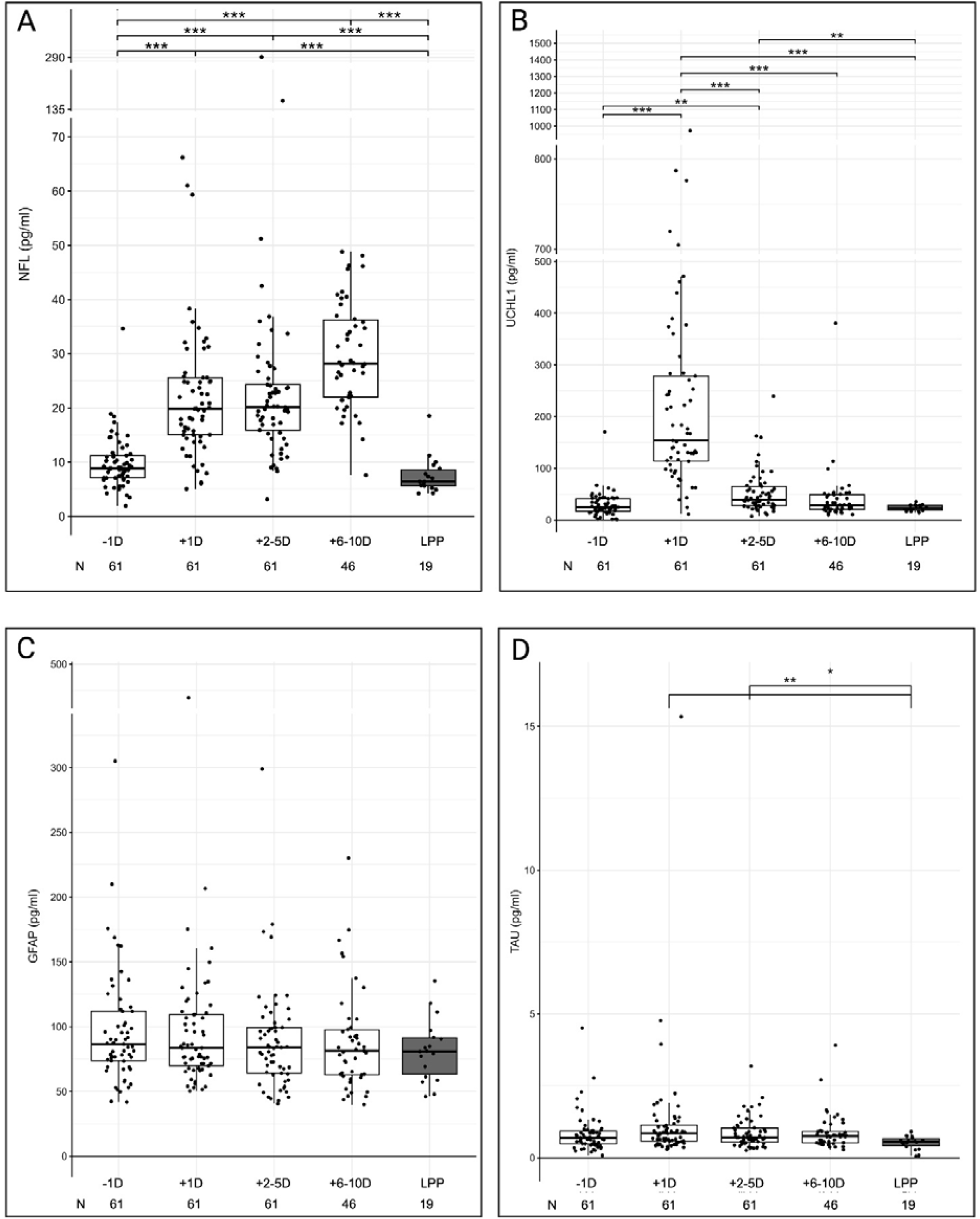
Pre- and post-partum serum levels of NfL, UCHL1, GFAP and TAU proteins. −1D: the day before partum; +1D: the day post-partum; +2-5D: 2-5 days post-partum; +6-10D: 6-10 days post-partum; LPP: late post-pregnancy, 20 −180 days post-partum. Boxes represent interquartile ranges (25th to 75th percentiles), and horizontal lines indicate median values. ** p < 0.001; *** p < 0.0001, (see test for statistical analysis). **2 A sNfL** = serum Neurofilaments Light **2 B sUCHL1** = serum Ubiquitin C-terminal Hydrolase L1 **2 C sGFAP** = serum Glial Fibrillary Acidic Protein **2 D sTAU** = serum Tubulin Associated Unit protein

### Levels of UCHL1 in Vaginal and C-section deliveries

A linear mixed-effects model was used to analyze the effect of delivery type on log-transformed sUCHL1 levels. The model, which accounted for time since delivery, parity, and intra-individual variability (as well as their interactions with time), revealed no statistically significant main effect of delivery type on UCHL1 levels over time (Table S1).

### Impact of parity on sUCHL1

In the same model, parity was found to be a time-independent factor influencing sUCHL1 levels. Similar to sNFL, sUCHL1 levels decreased with a higher number of previous pregnancies (b=−0.78, 95% CI [−0.96, −0.64], p=0.0202) (Table S1; Figure 3B).

**Figure 3.**
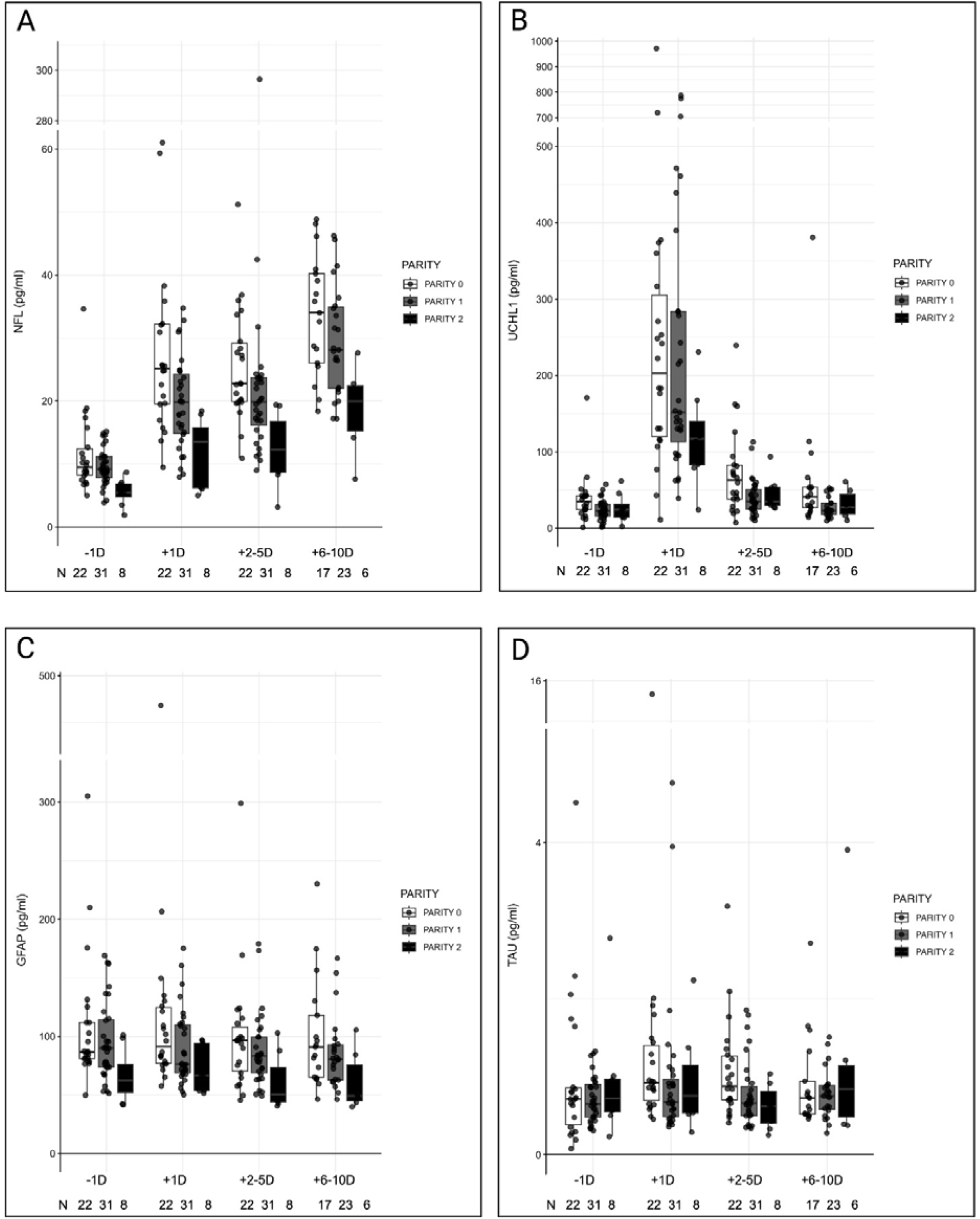
Impact of parity on pre- and post-partum sNfL, sUCHL1, sGFAP and sTAU levels. −1D: the day before partum; +1D: the day post-partum; +2-5D: 2-5 days post-partum; +6-10D: 6-10 days post-partum; Boxes represent interquartile ranges (25th to 75th percentiles), and horizontal lines indicate median values. ** p < 0.001; *** p < 0.0001, (see test for statistical analysis). **3 A sNfL** = serum Neurofilaments Light **3 B sUCHL1** = serum Ubiquitin C-terminal Hydrolase L1 **3 C sGFAP** = serum Glial Fibrillary Acidic Protein **3 D sTAU** = serum Tubulin Associated Unit protein

### sGFAP levels Over Time

sGFAP levels showed no significant changes across time points. Median values were 86.56 pg/ml (range: 41.77–305.05) at −1D, 83.51 pg/ml (range: 50.33–468.17) at +1D, 83.88 pg/ml (range: 40.58–298.90) at +2-5D, and 81.45 pg/ml (range: 39.85– 230.10) at +6-10D. All longitudinal time points were comparable to the LPP median of 81.08 pg/ml (range: 47.96–135.30) (Table 2, Figure 2C).

### sGFAP levels in Vaginal and C-section deliveries

A linear mixed-effects model with log-transformed sGFAP as the dependent variable showed no statistically significant main effect of delivery type on sGFAP levels over time. The model accounted for time since delivery, parity, type of birth, and their respective interactions with time, as well as intra-individual variability (Table S2).

### Impact of parity on sGFAP

Despite the temporal stability of sGFAP levels and their independence from delivery mode, a linear mixed-effects model demonstrated a significant inverse relationship with parity. The analysis, which used log-transformed sGFAP as the dependent variable and accounted for time since delivery, type of birth, and intra-individual variability (with their interactions with time), showed that sGFAP levels decreased as parity increased (b=−0.82, 95% CI [−0.94, −0.72], p=0.0065) (Table S2; Figure 3C).

### sTAU protein levels Over Time

Levels of sTAU did not differ significantly across the four longitudinal time points. However, a statistically significant increase was observed at +1D (median 0.56 pg/mL, range 0.06 – 0.91) compared to the LPP group (p=0.0089, Kruskal-Wallis test) (Table 2; Figure 2D).

### sTAU level in Vaginal and C-section Deliveries

A linear mixed-effects model with log-transformed sTAU as the dependent variable showed no statistically significant main effect of delivery type on sTAU levels over time. The model accounted for time from delivery, parity, and type of birth, along with their interactions with time, as well as intra-individual variability (p>0.05) (Table S3).

### Impact of Parity on sTAU

The linear mixed-effects model analysis demonstrated that the influence of parity on sTAU levels is time-dependent. Specifically, at the +2-5D time point, for every unit increase in parity, patients showed 0.74 times lower sTAU levels (p=0.04) (Table S3; Figure 3D). The model utilized log-transformed sTAU as the dependent variable and accounted for time from delivery, type of birth, parity, including its interaction with time, and intra-individual variability.

## Discussion

Our findings demonstrate that most HWwPP exhibit substantial increases in sNfL and sUCHL1 after delivery. sNfL elevation is detectable in late gestation, with significantly higher values at −1D versus LPP and N-PW groups. Childbirth triggers a sharp sNfL increase; levels double within hours postpartum and remain elevated for five days, peaking at +6-10D with median values sixfold higher than N-PW. While we cannot confirm this as the absolute peak, levels normalize within 40 days post-delivery. Birth mode (vaginal vs. C-section) does not differentially affect sNfL or other markers when analyzed using linear mixed-effects models accounting for time, parity, birth type, and individual variation.

The sNfL increase parallels sUCHL1 elevation, a deubiquitinating enzyme highly expressed in neurons and lactotroph cells of anterior pituitary.^15,16^ However, sUCHL1 increases are more intense and transient, peaking at +1D and returning to pre-partum levels by +6-10D.

Neuronal impact is further supported by elevated sTAU at +1D and +2-5D versus LPP. Conversely, GFAP, an astrocyte marker, shows no significant temporal changes. Consistent with previous studies, ^6,8,11^ postpartum sNfL elevation occurs in most but not all women. Notably, our findings suggest a potential explanation for the absence of an increase in some individuals: higher parity is associated with blunted sNfL elevation, with the magnitude diminishing as pregnancies increase. This inverse parity-biomarker association extends to sUCHL1, sGFAP, and sTAU. Parity’s impact on CNS structure remains understudied; it was unreported in prior postpartum sNfL studies^6-9,11^ and examined in only one neuroimaging study.17 The Rotterdam Study found increased cortical volume with higher parity in 2,835 women decades postpartum.^17^ While MRI studies focus predominantly on nulliparous women, converging evidence indicates pregnancy induces hormone-driven brain structural modifications in first-time mothers.^18^

Our study demonstrates a marked increase in neurological markers in the great majority of HWwPP without evident neurological pathology prior, during and after-pregnancy. A clinical implication of our findings is that elevated post-partum sNfL levels detected in wwMS^7-9^, or other CNS disorders^6,11^ during the early postpartum phase should not be solely attributed to new CNS damage.

Our study reflect a phenomenon, that exhibits the following characteristics: a) Elevated sNFL, indicative of CNS neuron involvement; b) Elevated sUCHL1, suggestive of involvement of neurons and/or lactotroph cells of the anterior pituitary; c) Triggered by parturition; d) Parity-dependent attenuation, as the intensity of the increase diminishes with higher parity; e) Self-limiting course, as it resolves within approximately 40 days; f) It does not result in any evident adverse health outcomes for the woman.

Our study cannot definitively determine whether this phenomenon represents a neuronal damage triggered by pregnancy and delivery or a physiological, harmless process reflecting pregnancy-induced neuroplasticity.^18^

Beyond neuroplasticity, in theory, biomarkers elevation could result from pregnancy-related distress, transient inflammation from rapid immunotolerance shifts, or non-CNS factors including age, variation of blood volume variation and/or Body Mass Index (BMI), or increased blood-brain-barrier (BBB) permeability. Alternatively, biomarkers may originate from the peripheral nervous system or other organs. Both stress and pregnancy are characterized by increased cortisol levels and activation of the hypothalamic-pituitary-adrenal axis. In non-pregnant individuals, stress induces structural changes in the CNS as an adaptation mechanism.^19^ However, our data do not support a role for stress in the observed biomarker changes. While vaginal delivery is considered more stressful than C-section delivery^20^, no differences in biomarkers were observed between the two groups in our study. Moreover, sNfL levels are not altered in stress-related mental disorders.^21^ Pregnancy causes a shift toward Th2 cytokines, followed by an inverse shift toward Th1 cytokines in the post-partum associated in wwMS with increased risk of relapses.^22^ In theory, this sequence might transiently induce inflammation and brain damage in HWwPP.

While age-related sNfL increases are well-established^14,23^, our cohort had a narrow age range (18-42 years) where increments are marginal in healthy controls.^23^ Moreover, women with parity ≥2, showing the smallest biomarker increases, were age-matched to those with parity 0-1.

While delivery causes acute reductions in blood volume and BMI, and increased BBB permeability occurs in animal pregnancy models^24^, these physiological changes cannot fully explain the magnitude and temporal patterns of biomarker alterations observed in this study, which differ in magnitude and temporal evolution.

It is plausible that sNfL originates from lesions of uterine or perineal nerves during parturition. However, our study did not reveal differences between vaginal and C-section delivery. Furthermore, the sharp increase of UCHL1, a protein primarily localized in the cytoplasm^15,16^, on the first day after delivery is difficult to explain by peripheral nerve injury.

While elevated serum biomarkers consistently indicate neuronal injury in adults^1-5^, our study demonstrate that sNfL and sUCHL1 can increase under healthy, non-pathological conditions such as the postpartum period in healthy women. Another physiological condition characterized by high sNfL levels occurs in healthy children, where sNfL decreases from birth to 10 years of age,^25^ a period characterized by intense cerebral neuroplasticity.

Pregnancy-induced neuroplasticity in nulliparous individuals has received increasing attention. Evidence from neuroimaging, hormonal, neuropsychological, and immunohistological studies in women and animals suggests neuroplasticity is the anatomical basis by which the CNS adapts to gestation and prepares for offspring care.

Our study cannot determine whether increased biomarkers originate from specific CNS areas or reflect diffuse brain structural modifications.^26,27^ These changes may reflect distinct events occurring at different times across diverse regions, supported by the differing timeframes of UCHL1 and sNfL increases. However, data from women and animal studies suggest most changes involve the cortex, hypothalamus, and pituitary gland, though some studies have highlighted hippocampal^28^ and striatal^29^ modifications.

Cortical gray matter reduction was initially demonstrated by MRI studies.^30^ Recent longitudinal MRI studies of first-time pregnant women consistently show gestational cortical gray matter volume reduction, followed by postpartum recovery.^31^ The same research group, utilizing the same cohort, recently reported that, unlike the cortex, the hypothalamus exhibits volumetric increases during pregnancy that recede postpartum in first-time mothers.^32^

Mouse neuroimaging reveals gray matter increases in hypothalamic subregions during late pregnancy and early postpartum, normalizing after weaning.^33^ Pregnancy-induced adult neurogenesis^34^ and extensive neurochemical, neuroendocrine, morphological, and gene expression alterations involve the medial Pre-Optic Area, Paraventricular Nucleus, and Supraoptic Nuclei. These regions regulate oxytocin and prolactin release, influencing uterine contraction, milk ejection, maternal behaviors, social bonding, and stress responses.

In pregnant women, the pituitary stalk widens and pituitary volume significantly expands to 120% of pre-pregnancy size.^35^ This results from placental estrogen-induced hyperplasia and hypertrophy of lactotrophs cells, which constitute 20–50% of the anterior pituitary. Maximum enlargement occurs 0–3 days postpartum, returning to normal within 6 months post-partum.^35^

While UCHL1 is expressed in all neurons, it is highly detected in lactotroph and gonadotroph cells of the mouse anterior pituitary.^16^ Since gonadotrophs are inhibited while lactotrophs increase during pregnancy, some postpartum sUCHL1 elevation, following placental elimination, may originate from lactotrophs, and some sNfL from pituitary stalk axons.

Assuming that the changes in serum biomarkers evidenced in our study reflect physiological neuroplasticity, we propose that the observed changes are due to pregnancy-induced neuroplasticity, leveraging the CNS’s inherent capacity for learning. Essentially, we suggest the CNS adapts during a woman’s first pregnancy, enabling more effective management of subsequent gestations.

In nulliparous women, pregnancy—likely via hormones or placental extracellular vesicles—induces the development and maintenance of new neurons within the CNS and lactotroph cells in the anterior pituitary gland (Figure 4). This process adapts the CNS to pregnancy, involving both modifications in hormone synthesis and psychological adjustments for offspring care.

**Figure 4.**
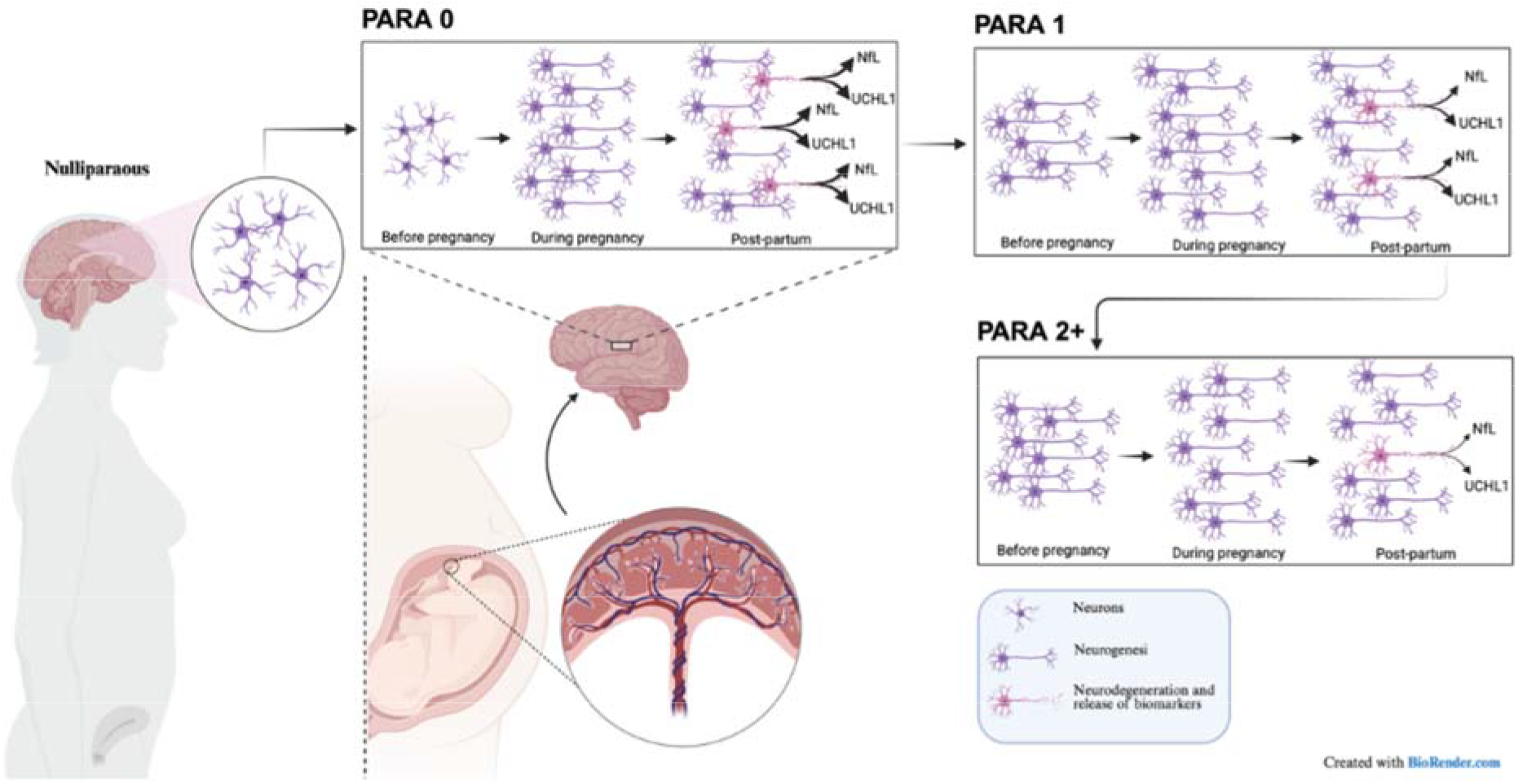
“Visual Representation of Pregnancy-Induced Neuroplasticity and the Impact of Subsequent Pregnancies. During a nulliparous pregnancy, the placenta, likely through hormonal influences, promotes the development and maintenance of new neurons and lactotroph cells. Upon parturition, the expulsion of the placenta removes the crucial support for these pregnancy-induced cells, leading to their demise and the subsequent release of biomarkers such as UCHL1, NfL, and TAU. However, a subset of these newly formed neurons and their axons persist, enduring through subsequent pregnancies. In a second pregnancy, this neuroplastic process reoccurs; however, the established neuronal structures from the preceding pregnancy result in reduced neoformation and subsequent degeneration of neurons. This phenomenon appears to become progressively refined and less pronounced in subsequent gestations. PARA 0 = first pregnancy; PARA 1 = second pregnancy; PARA 2+ = third or subsequent pregnancy “*Created in BioRender. Bertolotto, a. (2025) https://BioRender.com/kkzhx3s* “

Upon parturition, the expulsion of the placenta removes the support for these pregnancy-induced cells. These neurons subsequently die, releasing UCHL1, NfL, and TAU. However, some newly formed neurons and axons persist for use in a subsequent pregnancy.

During the second pregnancy, the phenomenon recurs, but the female body benefits from the neuronal structures already established. Consequently, the neoformation of neurons is reduced, as is their subsequent destruction. In subsequent gestations, this phenomenon is progressively refined to the point of being minimal or no longer discernible.

Our study presents limitations. While our data demonstrated a statistically significant difference, additional research is required to corroborate the influence of parity on serum biomarker levels, given that this study included a restricted number of women with parity ≥2. Moreover, our investigation is solely biochemical, lacking corresponding imaging data that would facilitate a more precise definition of brain regions involved in neuroplasticity.

Further studies are required to ascertain whether quantifying pre- and postpartum biomarker levels can aid in identifying women who develop postpartum pathologies, such as postpartum depression.^36^

## Acknowledgment

The authors extend their gratitude to Sabdi Valverde Namay for the preparation of Figure 4.

## Contributors

AB is the guarantor of the study and accepts full responsibility for the work and/or the conduct of the study, had access to the data and controlled the decision to publish. Study concept and design: AB. Acquisition of data: PV, CIB, SeM, SiM, CB. Analysis and interpretation of data: AB. Drafting of the manuscript: AB. Critical revision of the manuscript for important intellectual content: PV, CIB, SeM, SiM, CB, ADS, LM. Statistical analysis: PV, CIB, SeM. Study supervision: AB.

## Patient consent for publication

Not applicable.

## Ethics approval

This study involved human participants and was approved by the Ethical Committee of San Luigi Gonzaga University Hospital (approvals numbers 18390/2019 and 7262/2019. All participants provided written informed consent.

## Provenance and peer review

Not commissioned; externally peer-reviewed.

## Data availability statement

Data are available upon reasonable request. Aggregated de-identified participant data will be shared upon motivated written request to the corresponding author.

