## Supplemental table 1, 2 and 3 for "Serum Neurofilament Light Chain Increases in Healthy Postpartum: Is It Subclinical Brain Damage or Neuroplasticity?"

|  | | **sUCHL1**, pg/ml | | | |
| --- | --- | --- | --- | --- | --- |
| Time-points | | -1D | +1D | +2-5D | +6-10D |
| All pregnancies | Number | 61 | 61 | 61 | 46 |
|  | Median (range) | 24.51  (1.58 – 170.63) | 153.69  (11.25 – 971.81) | 39.10  (10.16 – 239.35) | 27.74  (9.91 – 380.76) |
| Vaginal delivery | Number | 28 | 28 | 28 | 19 |
|  | Median (range) | 26.147  (2.04 -170.63) | 156.02  (11.25-775.90) | 48.05  (7.39-239.35) | 30.59  (12.84-360.76) |
| C-section | Number | 33 | 33 | 33 | 27 |
|  | Median (range) | 23,43  (1.01-66.85) | 153.69  (39.18-971.81) | 33.37  (10.16-162.29) | 23.90  (9.91-113.54) |
| Parity 0 | Number | 22 | 22 | 22 | 17 |
|  | Median (range) | 35.14  (1.02 - 170.63) | 202.82  (129.27 - 971.81) | 62.98  (7.39 - 239.35) | 41.36  (14.54 - 380.76) |
| Parity 1 | Number | 31 | 31 | 31 | 23 |
|  | Median (range) | 22.80  (1.58 - 57.53) | 151.56  (39.18 - 786.98) | 35.16  (10.16 - 112.86) | 23.14  (9.91 - 52.70) |
| Parity ≥ 2 | Number | 8 | 8 | 8 | 6 |
|  | Median (range) | 24.09  (2.21 - 61.72) | 117.19  (24.07 - 230.71) | 35.02  (26.52 - 93.71) | 26.97  (10.38 - 60.91) |

**Supplemental Table 2**

|  | | **sGFAP**, pg/ml | | | |
| --- | --- | --- | --- | --- | --- |
| Time-points | | -1D | +1D | +2-5D | +6-10D |
| All pregnancies | Number | 61 | 61 | 61 | 46 |
|  | Median (range) | 86.56  (41.77 - 305.05) | 83.51  (50.33 - 468.17) | 83.884  (40.58 - 298.90) | 81.449  (39.85 - 230.17) |
| Vaginal delivery | Number | 28 | 28 | 28 | 19 |
|  | Median (range) | 87.21  (41.77 - 305.05) | 84.21  (51.57 - 468.17) | 89.368  (40.58 - 298.90) | 78.140  (39.85 - 230.14) |
| C-section | Number | 33 | 33 | 33 | 27 |
|  | Median (range) | 84.56  (42.27 - 209.88) | 76.78  (50.33 - 206.51) | 80.69  (42.74 - 169.28) | 82.42  (46.29 - 174.71) |
| Parity 0 | Number | 22 | 22 | 22 | 17 |
|  | Median (range) | 86.79  (49.86 - 305.05) | 91.38  (57.54 - 468.17) | 96.41  (45.55 - 298.90) | 90.96  (46.49 - 230.14) |
| Parity 1 | Number | 31 | 31 | 31 | 23 |
|  | Median (range) | 89.83  (51.40 - 168.87) | 76.26  (50.33 - 175.23) | 83.39  (49.20 - 178.99) | 80.46  (46.29 - 166.65) |
| Parity ≥ 2 | Number | 8 | 8 | 8 | 6 |
|  | Median (range) | 62.34  (41.77 - 101.13) | 66.75  (51.57 - 97.04) | 50.46  (40.58 - 102.89) | 49.28  (39.85 - 105.67) |

**Supplemental Table 3**

|  | | **sTAU**, pg/ml | | | |
| --- | --- | --- | --- | --- | --- |
| Time-points | | -1D | +1D | +2-5D | +6-10D |
| All pregnancies | Number | 61 | 61 | 61 | 46 |
|  | Median (range) | 0.69  (0.08 - 4.51) | 0.85  (0.29 - 15.34) | 0.71  (0.25 - 3.18) | 0.75  (0.28 - 3.91) |
| Vaginal delivery | Number | 28 | 28 | 28 | 19 |
|  | Median (range) | 0.72  (0.28 - 4.51) | 0.84  (0.46 - 3.95) | 0.65  (0.33 - 1.63) | 0.73  (0.45 - 3.91) |
| C-section | Number | 33 | 33 | 33 | 27 |
|  | Median (range) | 0.65  (0.08 - 2.29) | 0.87  (0.29 -15.34) | 0.77  (0.25 - 3.18) | 0.77  (0.28 - 2.71) |
| Parity 0 | Number | 22 | 22 | 22 | 17 |
|  | Median (range) | 0.72  (0.08 - 4.51) | 0.92  (0.46 - 15.34) | 0.88  (0.41 - 3.18) | 0.73  (0.46 - 2.71) |
| Parity 1 | Number | 31 | 31 | 31 | 23 |
|  | Median (range) | 0.65  (0.31 - 1.33) | 0.67  (0.36 - 4.77) | 0.66  (0.34 - 1.85) | 0.75  (0.28 - 1.51) |
| Parity ≥ 2 | Number | 8 | 8 | 8 | 6 |
|  | Median (range) | 0.72  (0.23 - 2.78) | 0.76  (0.29 - 2.24) | 0.62  (0.25 - 1.04) | 0.85  (0.38 - 3.91) |

**Supplemental Table 1**

**UHCL1 serum level pre- and post-partum: impact of vaginal and C-section delivery and of parity**

Abbreviations

sUCHL1 = serum Ubiquitin C-terminal Hydrolase L1; Time-points: -1D: the day before partum; +1D: the day post-partum; +2-5D: 2-5 days post-partum; +6-10D: 6-10 days post-partum; Parity 0 = first pregnancy; Parity 1 = second pregnancy; Parity ≥ 2 = third or fourth pregnancy

**Supplemental Table 2**

**GFAP serum level pre- and post-partum: impact of vaginal and C-section delivery and of parity**

Abbreviations: sGFAP = serum Glial Fibrillary Acidic Protein; Time-points: -1D: the day before partum; +1D: the day post-partum; +2-5D: 2-5 days post-partum; +6-10D: 6-10 days post-partum; Parity 0 = first pregnancy; Parity 1 = second pregnancy; Parity ≥ 2 = third or fourth pregnancy

**Supplemental Table 3**

**TAU serum level pre- and post-partum: impact of vaginal and C-section delivery and of parity**

Abbreviations: sTAU = serum Tubulin Associated Unit protein; Time-points: -1D: the day before partum; +1D: the day post-partum; +2-5D: 2-5 days post-partum; +6-10D: 6-10 days post-partum; Parity 0 = first pregnancy; Parity 1 = second pregnancy; Parity ≥ 2 = third or fourth pregnancy
